# Impact of COVID-19 on Antenatal Care Utilization and Health Provider Response in Tamale, Ghana: A Mixed-Method Study

**DOI:** 10.1101/2025.05.27.25327765

**Authors:** Azawutor Addae Eunice, Yankyera Owusu Benedict, Mensah Rudolph, Sarfo Bismark, Otupiri Easmon

## Abstract

**Background:** The COVID-19 pandemic disrupted essential maternal health services in low-resource settings, with uncertain impacts on antenatal care (ANC) utilization and delivery.

**Objective:** To evaluate the impact of COVID-19 on ANC service utilization in the Tamale Metropolitan Area, Northern Ghana, focusing on access barriers, healthcare provider experiences, and the feasibility of alternative delivery models.

**Design:** Facility-based, convergent mixed-methods cross-sectional study.

**Setting:** Four public health facilities in Tamale, Ghana.

**Participants:** 240 pregnant women aged 18–45 years receiving ANC and 25 maternal healthcare providers (midwives and nurses).

**Methods:** Structured surveys with women and five focus group discussions with providers were conducted. Secondary ANC attendance data from the District Health Information Management System (DHIMS2) were reviewed. Quantitative data were analyzed using descriptive statistics, chi-square tests, and logistic regression in Stata 16. Qualitative data were thematically analyzed.

**Results:** In 2020, antenatal care (ANC) attendance experienced a significant decline of 67.5%. Approximately 73.3% of women reported reduced visits, primarily due to fear of infection, transportation barriers (67.5%), or misinformation. Women who perceived a higher risk of infection at healthcare facilities were significantly more likely to decrease attendance (p<0.05). Healthcare providers reported challenges including staff shortages, increased workloads, and emotional stress, although improvements in infection control measures and shift systems were noted. Virtual care was utilized by 52.5% of women; however, 77.8% encountered internet-related challenges. No stock-outs of ANC medications were reported.

**Conclusion:** The COVID-19 pandemic exposed vulnerabilities in ANC delivery in Tamale, driven by access barriers and systemic constraints. Investments in digital health infrastructure, risk communication, and provider support are critical to enhance maternal healthcare resilience during public health crises.

## 1.0 INTRODUCTION

Maternal health remains a cornerstone of global public health and a critical marker of health system performance and social development. In low- and middle-income countries (LMICs), persistent structural barriers such as inadequate infrastructure, workforce shortages, and financial constraints continue to impede access to quality maternal care (1).

Antenatal care (ANC), as the initial point into the maternal healthcare continuum, is vital for reducing maternal and neonatal morbidity and mortality. Despite improvements in global ANC coverage, pregnancy and childbirth complications remain leading causes of preventable death worldwide. In 2023, an estimated 260,000 maternal deaths occurred globally, with 94% of them concentrated in LMICs where health system weaknesses and inequitable access to skilled care persist (2).

Sub-Saharan Africa and Southern Asia accounted for approximately 87% of these deaths, with Sub-Saharan Africa bearing the largest burden at around 70% (3). This stark disparity highlights the urgent need for strengthened health systems and targeted interventions in regions most affected by maternal mortality (4). The WHO reported approximately 178,000 maternal deaths in SSA in 2023, averaging 487 deaths per day (5).

In Ghana, maternal mortality remains a significant public health challenge. The maternal mortality ratio (MMR) was estimated at 319 deaths per 100,000 live births in 2023, significantly exceeding the Sustainable Development Goal target of fewer than 70 per 100,000 by 2030 (6). These figures reflect persistent gaps in maternal health services provision and utilization, particularly in under-resourced regions such as Northern Ghana. Access to comprehensive ANC, recommended by WHO as a minimum of eight contacts per pregnancy, remains limited due to systemic and socio-cultural barriers (7).

The COVID-19 pandemic, emerging in early 2020, introduced unprecedented shocks to health systems globally, disrupting essential services including maternal healthcare. Governments, including Ghana’s, implemented lockdowns, travel restrictions, and emergency health system restructuring to curb viral transmission. While necessary, these measures inadvertently constrained access to primary healthcare. ANC delivery was notably affected due to healthcare worker redeployment to COVID-19 duties, transportation restrictions, and patients’ fear of infection (8). Pregnant women, physiologically and immunologically vulnerable, faced increased risk of adverse outcomes from disrupted care (9).

Even before the pandemic, Ghana’s maternal healthcare system faced significant structural challenges: workforce shortages, inadequate infrastructure, and inequitable service distribution (10). These systemic limitations were especially acute in the Northern Region, where the health system has historically been under-resourced (10). These vulnerabilities, particularly in the Northern Region, were exacerbated during COVID-19. Pregnant women encountered reduced provider availability, appointment cancellations, limited digital health adoption, and logistical barriers (11). For healthcare professionals, service delivery was complicated by shortages of personal protective equipment (PPE), stringent infection prevention requirements, and patient hesitancy (11). These factors highlight the fragility and limited surge capacity of maternal health systems during public health emergencies.

This study investigates the impact of the COVID-19 pandemic on ANC services in the Tamale Metropolitan Area of Northern Ghana. Employing a mixed-methods approach, it explores pandemic-related changes in ANC utilization, barriers experienced by pregnant women, and adaptive strategies by maternal health providers. It further examines the role of telemedicine and mobile health (mHealth) technologies as continuity tools during crises. By documenting real- world experiences from a region disproportionately affected by systemic health inequities, this study contribute to the literature on health system resilience and informs context-specific maternal health policy (12).

Tamale, as the administrative, commercial, and healthcare center of Northern Ghana, offers a strategic setting for this investigation. Despite metropolitan status, structural barriers such as poverty, limited transportation infrastructure, and poor digital connectivity persist, and were further aggravated by the pandemic. (13). These conditions provide a unique opportunity to examine the intersection of systemic constraints and pandemic-related disruptions on ANC delivery. Across LMIC, lockdowns, infection fears, health workforce shortages, and shifting policy priorities reduced utilization of essential services during the pandemic (14). In Ghana, Ministry of Health’s infection control protocols and COVID-19 resource reallocations often came at the expense of routine maternal services, especially in under-resourced regions like Northern Ghana (15). Comparable evidence from Kenya, Nigeria, and Bangladesh reveals declines in ANC attendance, missed appointments, and delayed care initiation (16–18).

While quantitative research has documented reduced maternal service uptake during COVID-19, limited focus has been placed on healthcare providers’ experiences and service adaptations. This study addresses that gap by assessing how the pandemic affected ANC access, provider responses, and the use of digital health tools in Tamale. The findings aim to inform maternal health planning, enhance emergency preparedness, and promote equitable ANC access in future public health crises (13).

## 2.0 METHODS

### 2.1 Study Design

This facility-based cross-sectional study employed a convergent mixed-methods design to comprehensively assess the impact of the COVID-19 pandemic on antenatal care (ANC) services in the Tamale Metropolitan Area. The quantitative component involved administering a structured questionnaire to pregnant women (n = 240), capturing data on ANC utilization patterns and COVID-19-related disruptions. The qualitative component consisted of five focus group discussions (FGDs), each comprising five maternal healthcare providers (total n = 25), aimed at exploring operational challenges, provider experiences, and service adaptation strategies during the pandemic.

This design allowed for methodological triangulation, integrating quantitative and qualitative data to develop a nuanced understanding of service user experiences and healthcare provider perspectives. The use of a convergent design enhanced the validity and depth of findings by facilitating convergence and corroboration across data sources (19,20).

### 2.2 Study Setting

The study was conducted in the Tamale Metropolitan Area (TMA) of the Northern Region of Ghana. TMA is served by a network of public health facilities, including hospitals, health centres, and Community-based Health Planning and Services (CHPS) compounds. According to the 2021 Population and Housing Census, the total fertility rate in TMA is 2.8, which is lower than the regional average of 3.5, reflecting a diverse demographic landscape encompassing both urban and peri-urban communities with varying socio-economic profiles (21). Four facilities were purposively selected for the study based on ANC service utilization and geographic coverage: Bilpeila Health Centre, Vittin Health Centre, Kpanvo CHPS, and Lahagu CHPS. These sites reflect urban and peri-urban settings commonly accessed by pregnant women in the metropolis.

### 2.3 Study Population and Sampling

#### 2.3.1 Quantitative Component

The study targeted pregnant women aged 18–45 years who were currently receiving ANC services and had given birth at least once before March 2020, allowing for comparison between pre-pandemic and pandemic care experiences.

Sample size determination followed statistical principles to ensure adequate power. Using the Cochran formula for cross-sectional studies (Cochran, 1977).

Using the formula:

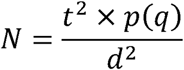

Where:

- **N** = Estimated sample size
- **t** = Confidence level (1.96 for 95%)
- **p** = 0.18 (estimated ANC service disruption prevalence based on preliminary studies)
- **q** = Probability of non-occurrence (1 – p = 0.82)
- **d** = Margin of error (0.05)

Substituting the values:

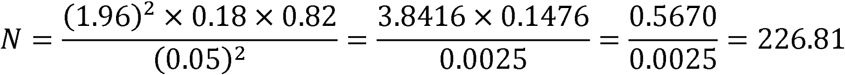

The calculation yielded a minimum sample of 226 participants. Accounting for a potential non-response rate of 6%, the final target sample was adjusted to **240 respondents**.

Systematic random sampling was employed to ensure representativeness. The list of ANC registrants at each facility served as the sampling frame. The total number of eligible clients was divided by the sample size to determine the sampling interval (k). The first participant was randomly selected, after which every kth individual was recruited until the sample size was achieved.

#### 2.3.2 Qualitative Component

The qualitative sample comprised maternal healthcare providers with ANC experience both before and during the pandemic, selected through purposive sampling to ensure informed perspectives on service adaptations (22). Five focus group discussions (FGDs) were conducted, each comprising five healthcare providers, resulting in a total of 25 participants. The sample size determination followed qualitative research standards, based on the principle of information saturation(23). During analysis, saturation was confirmed when no new themes emerged from the final FGD.

### 2.4 Data Collection Procedures

#### 2.4.1 Instrumentation

Quantitative data were collected using a structured questionnaire informed by global and national maternal health literature and adapted to the Ghanaian context. It captured socio-demographics, ANC attendance, perceived barriers, and pandemic-related experiences. The tool included both closed- and open-ended items and was pre-tested for validity and consistency at Bilpeila Health Centre, a demographically similar non-study site.

Qualitative data were gathered using a semi-structured FGD guide developed from study objectives and relevant literature. Key themes included workload, risk perception, infection control, communication, and service adaptations. The guide, developed in English, was orally translated during discussions where needed and piloted with non-participant providers to ensure clarity and relevance.

#### 2.4.2 Primary Data Collection

Three trained research assistants with public health backgrounds conducted the quantitative surveys under the principal investigator’s supervision during the post-lockdown recovery phase. COVID-19 safety protocols were strictly observed.

FGDs were facilitated by an experienced qualitative researcher in safe, ventilated spaces near the study sites. Sessions were conducted in English or translated as needed, audio-recorded with consent, and supplemented with field notes.

#### 2.4.3 Secondary Data Collection

Secondary data on ANC attendance from 2018 to 2021 were obtained from the District Health Information Management System (DHIMS2) (24)to examine longitudinal service utilization trends before, during, and after the COVID-19 peak. These data triangulated with primary survey findings to contextualize reported disruptions within broader service patterns.

### 2.5 Data Management and Analysis

Quantitative data were entered into a password-protected Excel database and analyzed using Stata version 16 (StataCorp LLC, College Station, TX). Descriptive statistics (frequencies, percentages, means, standard deviations) summarized participant characteristics and ANC utilization patterns. Associations between categorical variables were examined using Pearson’s chi-square tests, while binary logistic regression identified predictors of reduced ANC utilization during the pandemic, controlling for confounders. Statistical significance was set at p < 0.05, and model fit was assessed via the Hosmer-Lemeshow goodness-of-fit test.

Qualitative data from focus group discussions (FGDs) were transcribed verbatim within 48 hours and verified against recordings for accuracy. Transcripts were imported into NVivo 12 (QSR International) for manual thematic analysis following Braun and Clarke’s six-phase framework: data familiarization, code generation, theme development, reviewing, defining, and reporting (25). Two researchers independently conducted open, axial, and selective coding to identify themes, calculating inter-coder reliability with Cohen’s kappa. Discrepancies were resolved by consensus with a third researcher, and member checking was performed by sharing preliminary findings with participants to validate interpretations.

Integration of quantitative and qualitative results adhered to the triangulation protocol described by O’Cathain et al. (2010) (26), systematically comparing findings to identify convergence, complementarity, and divergence.

### 2.6 Ethical Considerations

Ethical approval was granted by the Committee on Human Research, Publications and Ethics (CHRPE) at Kwame Nkrumah University of Science and Technology (Approval No. CHRPE/AP/48722) and the Tamale Metropolitan Health Directorate. Written institutional permissions were obtained from all participating facilities. Participants received comprehensive information about the study’s purpose, procedures, risks, and benefits, and provided written informed consent. Confidentiality was strictly upheld through anonymized coding and secure data storage.

## 3.0 RESULTS

### 3.1 Quantitative Results

#### 3.1.1 Socio-Demographic Characteristics of Respondents

Most participants were aged 26–35 years (59.6%) and predominantly Muslim (86.2%). Educational attainment was generally low, with 33.3% having no formal education and 34.2% completing only basic education. Nearly all respondents were married (98.3%). Employment was reported by 62.1%, primarily in trading or business (61.7%). A significant proportion (49.6%) had 3–4 children. Most respondents (85%) belonged to the Mole-Dagbani ethnic group. Household income was generally low, with 47.9% earning below GHS 500 monthly. Regarding antenatal care, over half (52.5%) attended 2–3 times during the study period, while 34.2% met the recommended minimum of four visits (Table 1).

**Table 1:** Summary: Socio-Demographic Characteristics of Study Participants (N = 240)

| Demographic characteristics | Frequency (n=240) | Percentage (%) |
| --- | --- | --- |
| **Age group (years)** |  |  |
| 17 – 25 | 75 | 31.2 |
| 26 – 35 | 143 | 59.6 |
| 36 – 43 | 22 | 9.2 |
| **Religion** |  |  |
| Christian | 33 | 13.8 |
| Islam | 207 | 86.2 |
| **Educational level** |  |  |
| No formal education | 80 | 33.3 |
| Basic | 82 | 34.2 |
| Secondary/High school | 63 | 26.3 |
| Tertiary | 15 | 6.2 |
| **Marital status** |  |  |
| Never married | 1 | 0.4 |
| Married | 236 | 98.3 |
| Separated/divorced/widowed | 3 | 1.3 |
| **Employment status** |  |  |
| Unemployed | 91 | 37.9 |
| Employed | 149 | 62.1 |
| **Occupation, if employed** |  |  |
| Agriculture/farming | 21 | 14.1 |
| Business/trading | 92 | 61.7 |
| Civil/public servant | 14 | 9.4 |
| Entrepreneurship | 22 | 14.8 |
| **Parity** |  |  |
| 1-2 | 81 | 33.7 |
| 3-4 | 119 | 49.6 |
| ≥5 | 40 | 16.7 |
| **Ethnicity** |  |  |
| Akan/Ewe | 10 | 4.2 |
| Grusi | 6 | 2.5 |
| Mole-dagbani | 204 | 85.0 |
| Other | 20 | 8.3 |
| **Monthly income (GHC)** |  |  |
| Below 500 | 115 | 47.9 |
| 500 – 999 | 85 | 35.4 |
| 1000 – 2000 | 40 | 16.7 |
| **Number of ANC attendance** |  |  |
| 1 | 32 | 13.3 |
| 2 – 3 | 126 | 52.5 |
| ≥4 | 82 | 34.2 |

#### 3.1.2 Impact of COVID-19 on Antenatal Care Services

The COVID-19 pandemic significantly disrupted ANC service utilization (Table 2). A majority (73.3%) of pregnant women reported fear of infection as a barrier to accessing ANC. Scheduled visits declined for 62.5% of respondents, and overall ANC attendance dropped for 67.5%, with 45% missing at least one appointment. While 57.1% continued in-person visits, 27.5% relied on virtual services and 15.4% used a hybrid model. Perceptions of provider availability varied: 37.9% were uncertain, 34.6% felt there was adequate staffing, and 27.5% perceived provider shortages. Although 61.7% reported that health facilities remained operational, 52.9% experienced transportation challenges.

**Table 2:**
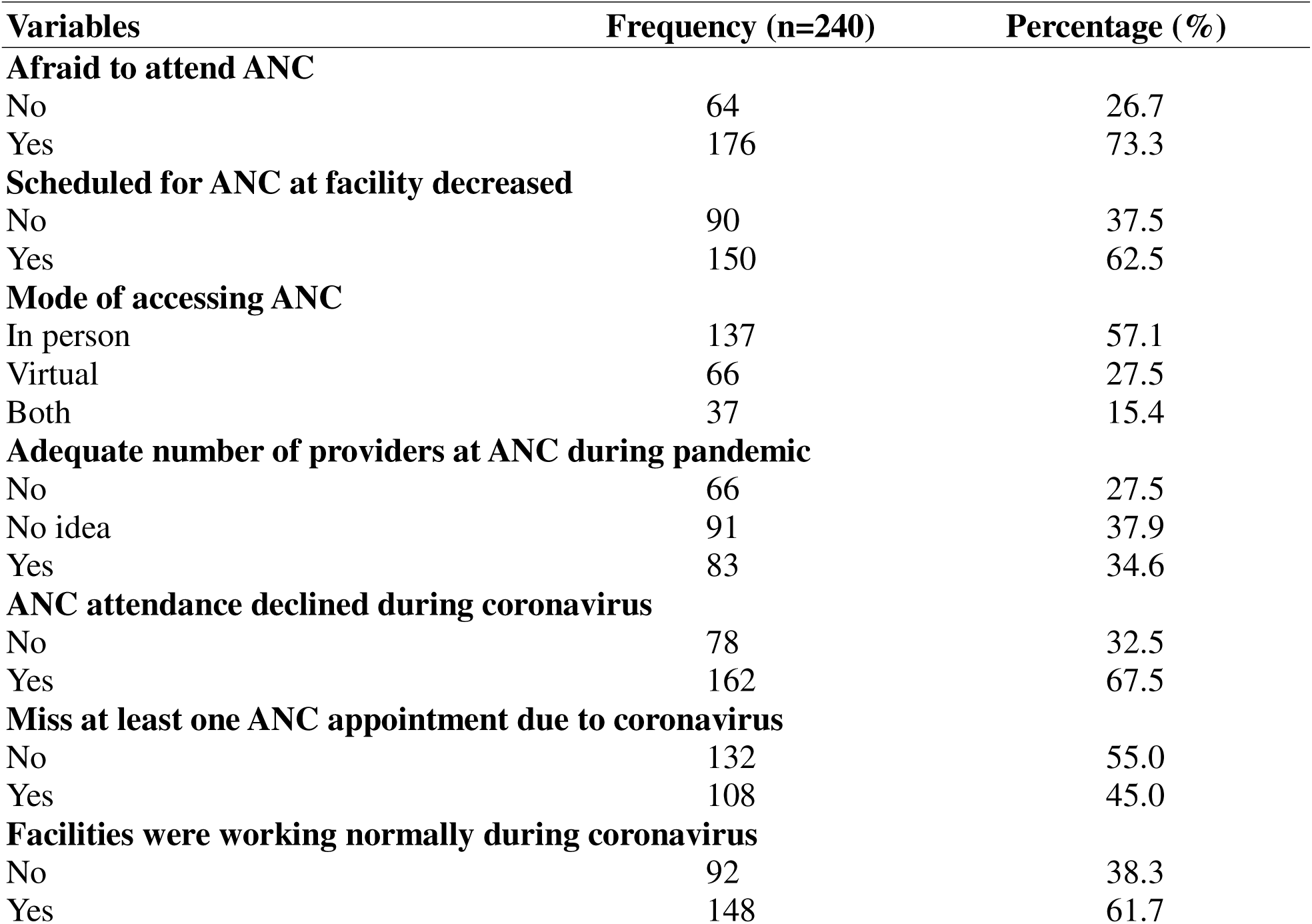
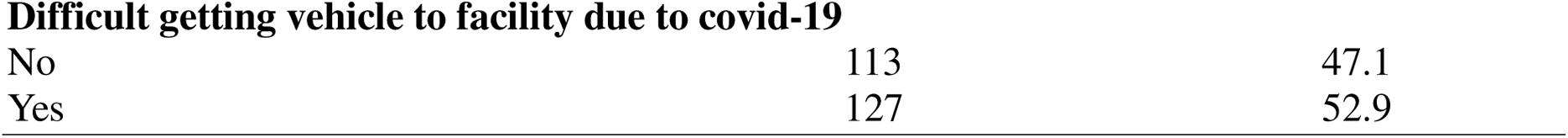
Influence of COVID-19 on Antenatal Care Utilization and Service Delivery in Tamale, Ghana.

| **Variables** | **Frequency (n=240)** | **Percentage (%)** |
| --- | --- | --- |
| **Afraid to attend ANC** |  |  |
| No | 64 | 26.7 |
| Yes | 176 | 73.3 |
| **Scheduled for ANC at facility decreased** |  |  |
| No | 90 | 37.5 |
| Yes | 150 | 62.5 |
| **Mode of accessing ANC** |  |  |
| In person | 137 | 57.1 |
| Virtual | 66 | 27.5 |
| Both | 37 | 15.4 |
| **Adequate number of providers at ANC during pandemic** |  |  |
| No | 66 | 27.5 |
| No idea | 91 | 37.9 |
| Yes | 83 | 34.6 |
| **ANC attendance declined during coronavirus** |  |  |
| No | 78 | 32.5 |
| Yes | 162 | 67.5 |
| **Miss at least one ANC appointment due to coronavirus** |  |  |
| No | 132 | 55.0 |
| Yes | 108 | 45.0 |
| **Facilities were working normally during coronavirus** |  |  |
| No | 92 | 38.3 |
| Yes | 148 | 61.7 |
| Difficult getting vehicle to facility due to covid-19 |  |  |
| No | 113 | 47.1 |
| Yes | 127 | 52.9 |

These findings are further corroborated by administrative service data. Figure 1 illustrates a substantial drop in ANC attendance in the Tamale Metropolitan Area, declining from 119,140 visits in 2019 to 94,744 in 2020, the height of pandemic-related restrictions, followed by a rebound to 116,467 visits in 2021. Similarly, Figure 2 highlights consistent patterns across selected health facilities, showing a marked reduction in ANC utilization during 2020. These data underscore the extent to which the COVID-19 pandemic disrupted maternal healthcare access, both at the community and facility levels.

**Figure 1:**
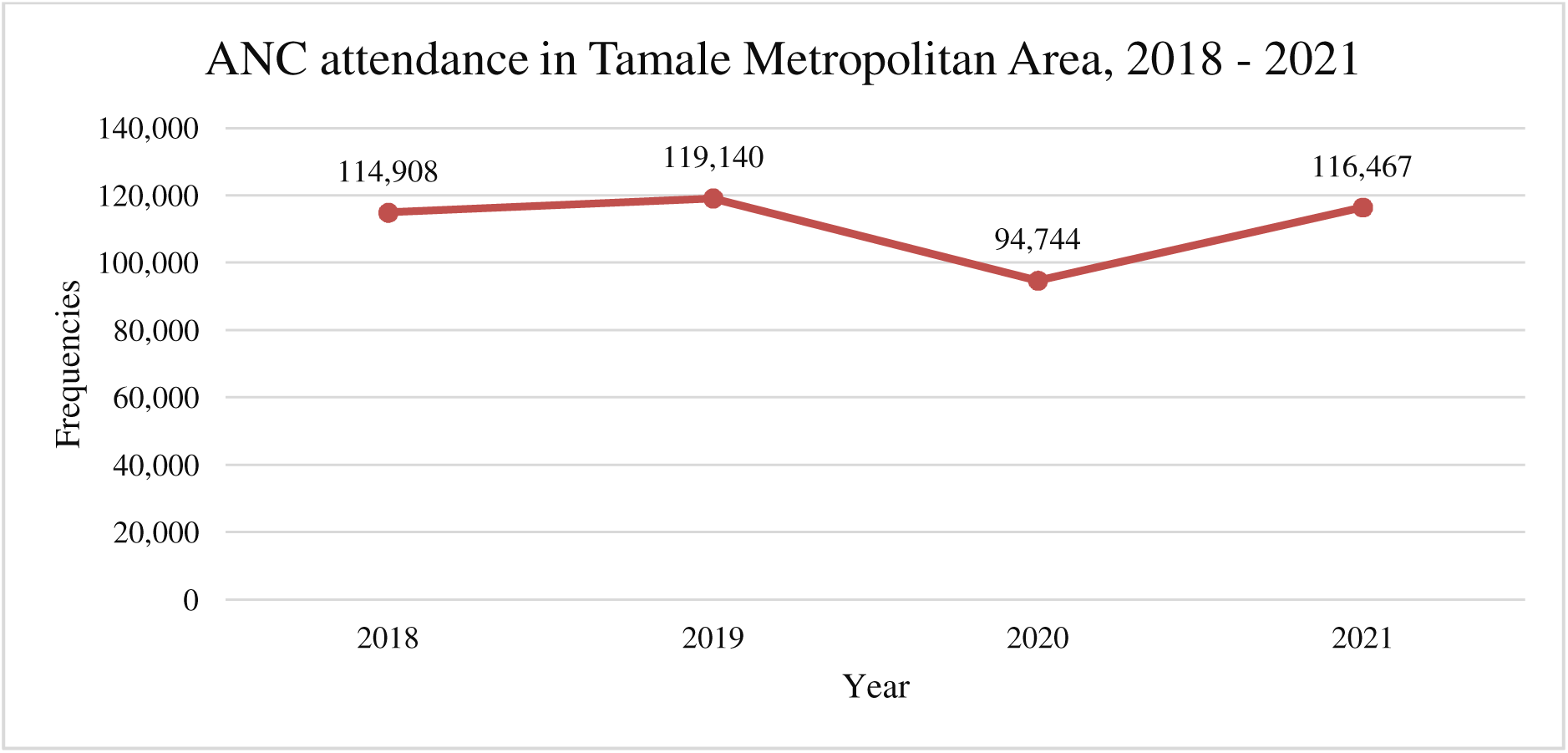
ANC attendance among pregnant women in Tamale Metropolitan Area, 2018-2021

**Figure 2:**
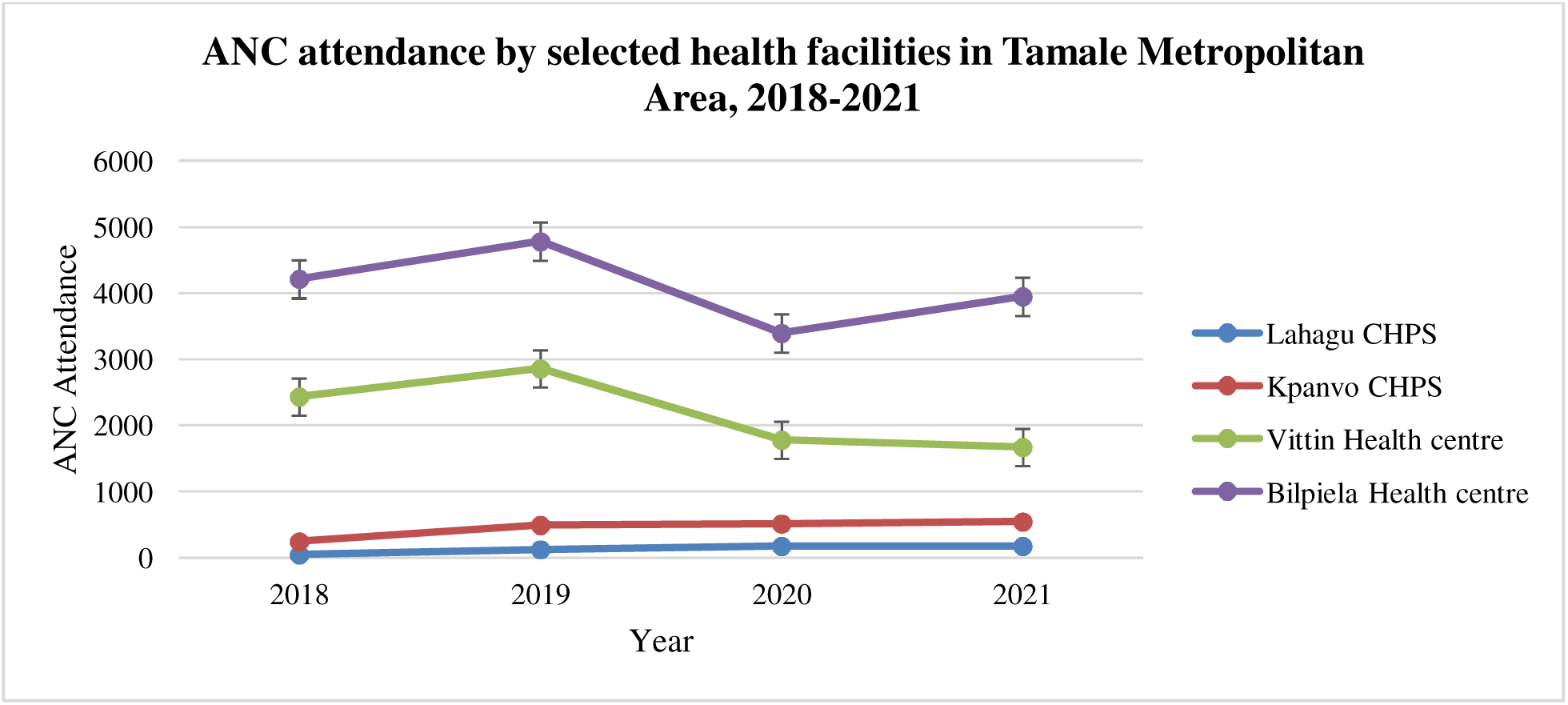
ANC attendance by selected health facilities in Tamale Metropolitan Area

#### 3.1.3 Accessibility to Antenatal Care Services During the Pandemic

Approximately 70% of respondents experienced delays in seeking ANC due to lockdowns and movement restrictions (Table 3). While 57.5% attended ANC in their first trimester, 41.3% began in the second trimester. Health complications were reported by 36.7% of participants. Despite these challenges, nearly all women completed iron and folic acid supplementation (96.2%) and sulphadoxine pyrimethamine prophylaxis (97.1%). Overall, 69.6% reported reduced ANC visits, 66.7% cited limited-service availability, and 63.3% experienced transport barriers.

**Table 3:**
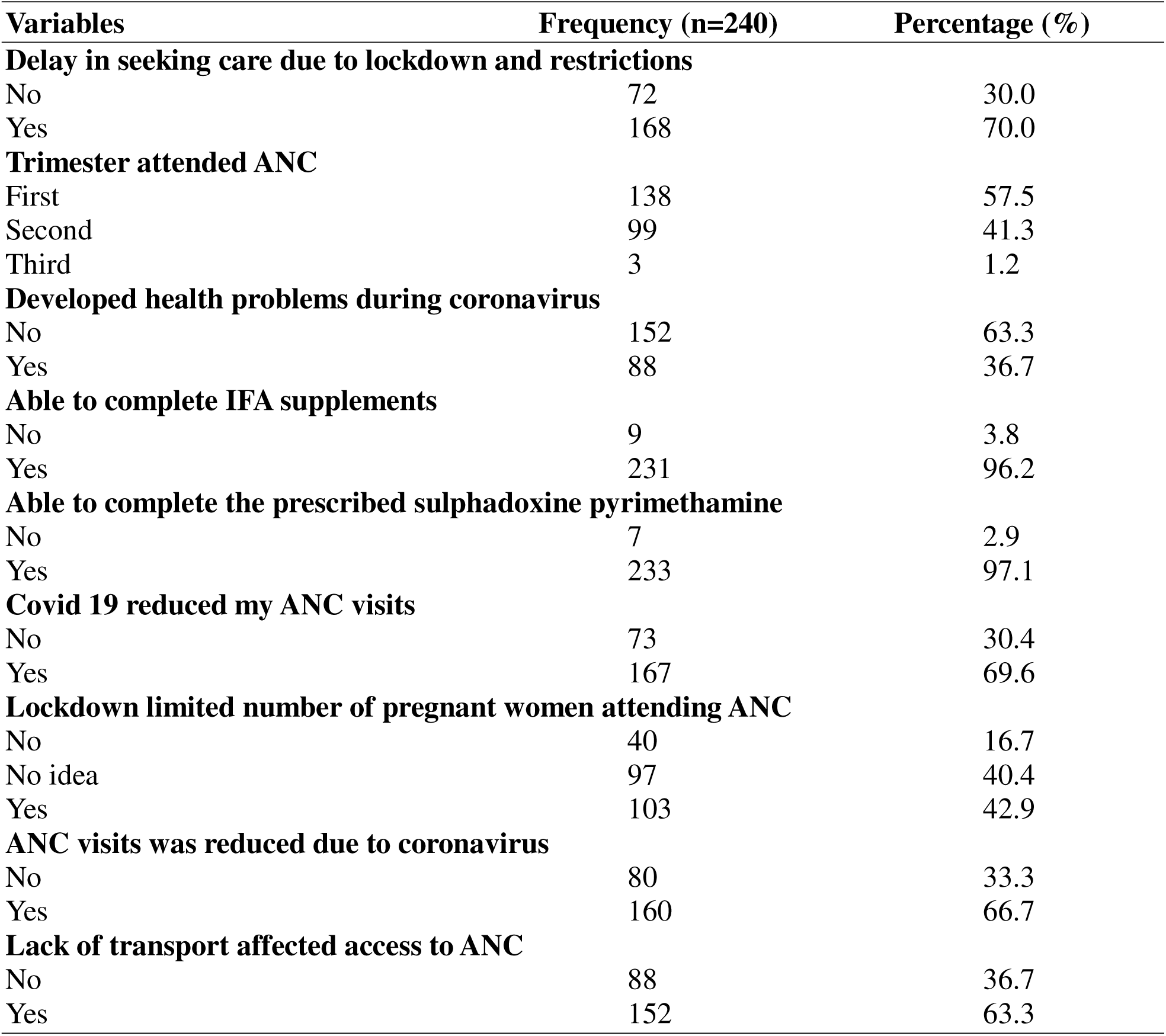
Accessibility to antenatal care among pregnant mothers during COVID-19 Variables Frequency (n=240) Percentage (%)

| **Variables** | **Frequency (n=240)** | **Percentage (%)** |
| --- | --- | --- |
| **Delay in seeking care due to lockdown and restrictions** |  |  |
| No | 72 | 30.0 |
| Yes | 168 | 70.0 |
| **Trimester attended ANC** |  |  |
| First | 138 | 57.5 |
| Second | 99 | 41.3 |
| Third | 3 | 1.2 |
| **Developed health problems during coronavirus** |  |  |
| No | 152 | 63.3 |
| Yes | 88 | 36.7 |
| **Able to complete IFA supplements** |  |  |
| No | 9 | 3.8 |
| Yes | 231 | 96.2 |
| **Able to complete the prescribed sulphadoxine pyrimethamine** |  |  |
| No | 7 | 2.9 |
| Yes | 233 | 97.1 |
| **Covid 19 reduced my ANC visits** |  |  |
| No | 73 | 30.4 |
| Yes | 167 | 69.6 |
| **Lockdown limited number of pregnant women attending ANC** |  |  |
| No | 40 | 16.7 |
| No idea | 97 | 40.4 |
| Yes | 103 | 42.9 |
| **ANC visits was reduced due to coronavirus** |  |  |
| No | 80 | 33.3 |
| Yes | 160 | 66.7 |
| **Lack of transport affected access to ANC** |  |  |
| No | 88 | 36.7 |
| Yes | 152 | 63.3 |

#### 3.1.4 Perception of Coronavirus on Antenatal Care Services

Majority of the respondents (75%) believed that COVID-19 disrupted their connection with maternal health providers (Table 4), while 63.8% felt the pandemic compromised the quality of maternal childcare. Emotional and physical support declined by 79.2%, and 77.5% reported heightened psychological stress. About 52.5% accessed virtual ANC, with 45% using online consultations; however, 77.8% faced internet connectivity challenges. Additionally, 68.3% experienced contact time with their providers, and 44.6% reported increased hospitalizations due to unscheduled ANC visits.

**Table 4:** Perception of Coronavirus on ANC.

| **Variables** | **Frequency (n=240)** | **Percentage (%)** |
| --- | --- | --- |
| **Coronavirus disconnected pregnant women from health providers** |  |  |
| No | 60 | 25.0 |
| Yes | 180 | 75.0 |
| **Coronavirus compromised quality maternal and child care** |  |  |
| No | 87 | 36.2 |
| Yes | 153 | 63.8 |
| **Coronavirus reduced emotional and physical support** |  |  |
| No | 50 | 20.8 |
| Yes | 190 | 79.2 |
| **Covid increased caesarean section delivery** |  |  |
| No | 162 | 67.5 |
| Yes | 78 | 32.5 |
| **Increased emotional and psychological stress during covid-19** |  |  |
| No | 54 | 22.5 |
| Yes | 186 | 77.5 |
| **Increased virtual ANC care during coronavirus** |  |  |
| No | 114 | 47.5 |
| Yes | 126 | 52.5 |
| **There were increase hospitalization due to unscheduled attendance** |  |  |
| No | 133 | 55.4 |
| Yes | 107 | 44.6 |
| **Use virtual consultation during coronavirus** |  |  |
| No | 132 | 55.0 |
| Yes | 108 | 45.0 |
| **If yes, did you encounter internet related issues** |  |  |
| No | 24 | 22.2 |
| Yes | 84 | 77.8 |
| **Reduction in contact time during coronavirus pandemic** |  |  |
| No | 76 | 31.7 |
| Yes | 164 | 68.3 |

### 3.8 Qualitative Results

#### 3.8.1 Background Characteristics of Participants

Twenty-five maternal healthcare providers participated in five FGDs. Most were aged 31–40 years (60%), female (72%) and married (96%). Participants were from four facilities: with Vittin Health Centre contributing the highest proportion (28%). The majority were midwives (80%), and Islam was the predominant religion (80%). In terms of experience, 40% had over eleven years of practice, while 32% had 1-5 years and 28% had 6-10 years. Just over half (52%) had 1-2 children (Table 5).

**Table 5:** Sociodemographic Characteristics.

| **Variables** | **Frequency (n=25)** | **Percentage (%)** |
| --- | --- | --- |
| ***Age group (years)*** |  |  |
| 21 – 30 | 6 | 24 |
| 31 – 40 | 15 | 60 |
| ≥41 | 4 | 16 |
| ***Gender*** |  |  |
| Male | 7 | 28 |
| Female | 18 | 72 |
| ***Marital status*** |  |  |
| Never married | 1 | 4 |
| Married | 24 | 96 |
| ***Facility*** |  |  |
| Kpanvo CHPS | 6 | 24 |
| Vittin health centre | 7 | 28 |
| Bilpiela health centre | 6 | 24 |
| Lahagu CHPS | 6 | 24 |
| ***Type of profession*** |  |  |
| Midwife | 20 | 80 |
| Nurse | 3 | 12 |
| Community health officer | 1 | 4 |
| Disease control officer | 1 | 4 |
| **Religion** |  |  |
| Islam | 20 | 80 |
| Christian | 5 | 20 |
| **Parity** |  |  |
| 0 | 3 | 12 |
| 1 – 2 | 13 | 52 |
| ≥3 | 9 | 36 |
| **Work experience** |  |  |
| 1 – 5 | 8 | 32 |
| 6 – 10 | 7 | 28 |
| ≥11 | 10 | 40 |

#### 3.8.2 Perceived Impact of COVID-19 on ANC Attendance

Thematic analysis of focus group discussions (FGDs) with maternal healthcare providers revealed seven key themes related to the impact of COVID-19 on antenatal care (ANC) services in the Tamale Metropolitan Area. The analysis, guided by the Braun & Clarke framework and supported by NVivo 12 software, yielded the following themes:

#### 3.1.1 Decreased ANC Utilization and Fear of Infection

Most providers reported a significant decline in ANC attendance during the pandemic, attributing it to widespread fear among pregnant women of contracting COVID-19 at health facilities.

> “COVID has really affected ANC attendance in the facility. Mothers felt that due to physical distancing, they were likely to contract the disease when visiting the facility.” (Midwife, anonymous)

> “Mothers heard that coronavirus spreads through droplets. So, when they come for ANC and are clustered, there are higher chances of infection.” (Nurse, anonymous)

#### 3.1.2 Enhanced Infection Prevention Measures

Participants highlighted improvements in hygiene practices, including handwashing and mask use, which were intensified during the pandemic.

> “Coronavirus increased handwashing among health workers and clients. Provisions were made and equipment were available at vantage points for immediate use.” (Midwife, anonymous)

#### 3.1.3 Modifications to Service Delivery

Facilities adapted by introducing shift systems and enforcing physical distancing. These changes were designed to minimize exposure while maintaining essential services.

> “We introduced a shift system so that one group of clients came on a particular day and another group came the next day. It reduced contact hours and helped protect both staff and clients.” (Midwife, anonymous)

> “Health workers were cautious. We strictly adhered to protocols—handwashing, PPEs, masks. Clients were also made to wear masks before entry.” (Staff nurse, anonymous)

#### 3.1.4 Absence of Digital or Virtual Consultations

Despite global trends toward digital health, none of the facilities reported implementing virtual ANC services. Barriers included poor internet access, client illiteracy, and financial limitations.

> “There were no forms of online or virtual consultations. Most clients could not be reached and had no access to internet or smartphones.” (Midwife, anonymous)

#### 3.1.5 Maternal Health Outcomes and Missed Preventive Interventions

Providers observed an increase in complications among pregnant women, particularly due to missed doses of intermittent preventive treatment for malaria (IPTp).

> “Pregnant women developed health problems during the pandemic. They weren’t coming for fansidar, so we saw more cases of complicated malaria.” (Midwife, anonymous)

#### 3.1.6 Availability of Health Commodities and Medicines

Contrary to expectations, there were no stockouts of essential maternal health commodities. Respondents attributed this to donations from government and non-governmental organizations.

> “There were no stockouts. The facilities even had more drugs and materials due to support from donors.” (Nurse, anonymous)

#### 3.1.7 Continuity of ANC Services

Despite challenges, providers emphasized that all essential ANC services continued during the pandemic, albeit with caution and modifications to ensure safety.

> “Nothing was left out. Every service was provided as usual, but we considered every client as potentially positive and took precautions.” (Midwife, anonymous)

## 4.0 DISCUSSION

### 5.1 Effects of COVID-19 on Antenatal Care Services

The COVID-19 pandemic disrupted global health systems, with maternal health services among the most affected sectors. This study corroborates a growing body of evidence demonstrating that fear of SARS-CoV2 infection significantly reduced antenatal care (ANC) attendance in low-resource settings such as Tamale, Ghana. Pregnant women reported skipping scheduled appointments and avoiding health facilities primarily due to concerns over virus exposure, a phenomenon documented extensively in multiple contexts (27–29).

In high-income countries, rapid adaptation included the deployment of virtual ANC platforms; for example, in Australia, telehealth interventions effectively halved in-person visits without compromising pregnancy outcomes (30,31). However, our findings emphasized such innovations remain largely inaccessible in many low- and middle-income countries (LMICs) due to infrastructural and socio-economic barriers. Poor internet connectivity, pervasive digital illiteracy, and limited financial resources significantly impede equitable adoption of telemedicine solutions (32,33). This digital divide underscores an urgent imperative to develop contextually appropriate, scalable digital maternal health interventions tailored to the realities of LMIC populations.

Our results align with reports from Saudi Arabia, where approximately one-third of pregnant women missed at least one ANC visit during the pandemic (34). Given the critical role of timely and consistent ANC in mitigating maternal and neonatal morbidity and mortality (35), such disruptions constitute a serious public health challenge. Moreover, physical access barriers compounded the problem: mobility restrictions and lockdown measures impeded transportation, further reducing service utilization. Similar observations have been documented in Ethiopia, where transport difficulties were a significant contributor to reduced ANC uptake during COVID-19 lockdowns (36).

Together, these findings highlight the multifaceted nature of pandemic-related barriers to maternal care, spanning psychosocial fears, systemic infrastructure deficits, and socioeconomic constraints. Addressing these requires integrated policy responses that not only restore pre-pandemic service levels but also fortify health system resilience against future shocks.

### 5.2 Accessibility to Antenatal Care During the Pandemic

The COVID-19 pandemic significantly curtailed access to antenatal care (ANC), particularly during the early months of the outbreak in 2020. In the Tamale Metropolitan Area, this study documented a marked decline in ANC attendance, coinciding with national lockdowns and heightened fear of infection. Similar reductions in service uptake have been observed globally; for example, Goyal et al. (2021) reported a 45.1% decrease in ANC utilization in India during the first wave of the pandemic (37).

Delayed initiation of ANC, frequently beginning in the second trimester, was commonly attributed to travel restrictions, facility closures, and psychological distress. These findings reflect global trends where pandemic-related disruptions led to delays in routine maternal care, exacerbating maternal and fetal health risks. Evidence from India suggests that such delays were associated with increased maternal complications and heightened levels of anxiety and depression among pregnant women (38).

Despite these challenges, most participants in this study reported completing essential interventions such as iron and folic acid (IFA) supplementation and sulphadoxine-pyrimethamine prophylaxis. This suggests that while access to routine checkups was compromised, some core components of ANC delivery remained resilient likely due to prioritization of these services by providers and their availability through community-based outlets.

However, the accessibility of ANC was undermined by a triad of interrelated barriers: economic hardship due to job losses or reduced income, transportation limitations, and inadequate access to trustworthy health information. These factors compounded existing inequities, particularly for women in underserved or peri-urban communities. The pandemic thus not only disrupted service delivery but also intensified social determinants of health, increasing the risk of adverse maternal and neonatal outcomes in already vulnerable populations.

### 5.3 Health professional’s perception on influence of coronavirus on antenatal care services

The study revealed that there was a disconnection between health staff and pregnant women due to the coronavirus pandemic as well as participants claimed coronavirus compromised quality maternal and childcare. In relation to the above finding, studies revealed that coronavirus pandemic had negative repercussions on the provision of antenatal care in multiple ways which include decreased psychological and physical support for women and compromised care standards, and increased exposure to medically unjustified caesarean section (39). Relatedly, a facility-based cross-sectional study in Sri Lanka also revealed that nearly half of women missed their routine antenatal clinic visits and the quality antenatal care has been compromised due to the coronavirus pandemic (40).

More than three-fourth (79.2%) claimed coronavirus reduced emotional and physical support for pregnant women and nearly one-third (32.5%) indicated coronavirus increased caesarean section delivery. Also, more than three-fourth (77.5%) had increased emotional and psychological stress during coronavirus. Similarly, a cross-sectional study indicated that pregnant mothers experienced increased psychological and emotional stress, as well as work challenges during the current corovirus pandemic due to receiving poor emotional and physical support (41). This is because, pregnant mothers were not utilizing antenatal care services during the coronavirus pandemic to receive pregnancy related counselling. Hence, the emotional and psychological support pregnant women receive from health providers was absent.

More than half of health workers tried the online/virtual antenatal care during coronavirus and nearly half (44.6%) claimed that there was increased hospitalization among pregnant women due to unscheduled antenatal attendance. Research conducted in the United Kingdom showed that coronavirus restrictions to prevent the spread of COVID-19 also reduced face-to-face interactions with healthcare professionals for maternity care. This meant that pregnant women failed to attend their antenatal visits or receive support in the postpartum period. Women were more uncertain about pregnancy and birth due to a lack of information on COVID-19 and pregnancy (42), and this led to health providers resulting to use online mode of healthcare accessibility. Among those who tried using the online/virtual form of consultation, more than three-fourth (77.8%) had issues with the internet connections. A UK systematic review and meta-analysis found a notable reduction in antenatal care services and unscheduled visits per week. Conversely, it reported an increase in virtual antenatal care and hospitalisations of unscheduled attendees (43). In Sri Lanka, a facility-based study revealed that maternal health providers specifically the midwives (90.3%) were the commonest source of information. However, internet access for online/virtual consultation was poor and this affected the quality of antenatal care pregnant mothers were receiving (44).

Also, coronavirus had significant impact on the timing of receiving care from health providers. Research revealed that more than two-thirds (68.3%) claimed that coronavirus pandemic reduced the contact time between pregnant women and health providers. The finding corroborates and affirms a research study in Kenya. In the qualitative study it was revealed that mothers feared attending hospitals for antenatal care because of the possibility of contracting COVID-19 and for those who attended, there was less contact time between providers and clients (45)

## 5.0 Implications of the Study

This study’s findings highlight critical public health implications for maternal healthcare delivery and pandemic preparedness in low- and middle-income countries (LMICs). The significant decline in antenatal care (ANC) utilization driven by fear of infection, transportation barriers, and absence of virtual alternatives reveals the urgent need to strengthen health system resilience to maintain essential services during crises. The lack of teleconsultation options highlights pervasive digital inequities, calling for investments in affordable, accessible digital infrastructure and literacy. Reported health complications due to reduced ANC visits emphasize the importance of adaptable service models, such as mobile clinics and community outreach. Notably, improved infection prevention and control (IPC) practices during the pandemic, including enhanced hygiene and PPE usage, should be institutionalized beyond emergency contexts. Finally, the psychological toll on both patients and providers signals a pressing need to integrate occupational and mental health support into maternal health systems.

## 6.0 Study Limitations

This study offers important insights into the impact of COVID-19 on antenatal care in Northern Ghana; however, several limitations must be noted. The findings, drawn from selected facilities in the Tamale Metropolitan Area, may not be generalizable to rural or other regions. Potential recall and social desirability biases could have influenced participants’ responses, particularly regarding events from the early phase of the pandemic. Although a significant proportion (62.5%) reported reduced ANC scheduled visits, the cross-sectional design limits causal interpretation, and facility-based data may not capture community-level trends or broader socioeconomic influences. Lastly, the absence of a longitudinal or pre/post-pandemic comparison restricts the ability to assess changes in service utilization over time.

## 7.0 Conclusion and Recommendations

This study confirms that the COVID-19 pandemic significantly disrupted antenatal care (ANC) services in Northern Ghana, leading to reduced attendance, missed appointments, and operational challenges for healthcare providers. While essential components such as iron and folic acid supplementation were largely maintained, the disruptions highlight the fragility of maternal health systems during crises. To enhance resilience and continuity of care, we recommend the development of a national telemedicine strategy tailored to ANC delivery, integration of emergency preparedness protocols into routine maternal health services, and strengthened public health communication to counter fear and misinformation. At the facility level, flexible scheduling systems and sustained infection prevention and control (IPC) measures should be institutionalized. Community engagement must be reinforced through the deployment of community health workers for follow-up and education, alongside targeted health literacy campaigns. Finally, future research should explore the feasibility and equity implications of virtual ANC models and assess the long-term maternal and neonatal impacts of pandemic-related service disruptions. Implementing these strategies will strengthen maternal healthcare systems and improve their responsiveness in the face of future public health emergencies.

## Data Availability

Data available upon request from the corresponding author.

## Supporting information

Ethical Approval

information Sheet and consent form

Qualitative Analysis

## Data Availability

All relevant data supporting the findings of this study are included in the manuscript and its supplementary files. Individual level data can be made available upon reasonable request from the corresponding author, subject to approval by the Committee on Human Research, Publications and Ethics (CHRPE), KNUST/Komfo Anokye Teaching Hospital.

