## Supplementary material for "Impact of COVID-19 on Antenatal Care Utilization and Health Provider Response in Tamale, Ghana: A Mixed-Method Study": Ethical Approval

**Kwame Nkrumah**
University of Science
and Technology, Kumasi

College of Health Sciences
SCHOOL OF MEDICINE AND DENTISTRY

COMMITTEE ON HUMAN RESEARCH, PUBLICATION AND ETHICS

Our Ref: CHRPE/AP/487/22

16th August, 2022

Ms. Eunice Addae Azawutor
Department of Population
Family and Reproductive Health
School of Public Health
KNUST-KUMASI

Dear Madam,

**LETTER OF APPROVAL**

***Protocol Title:*** ***Effect of Covid-19 on Antenatal Care Services in the Tamale Metropolitan Area, Northern Region, Ghana.***

***Proposed Site:*** ***Vittin Health Centre, Bilpeila Health Centre, Kpanvo CHPS and Kotingle CHPS.***

***Sponsor:*** ***Self Sponsored.***

Your submission to the Committee on Human Research, Publications, and Ethics on the above-named protocol refer.

The Committee reviewed the following documents:

- A notification letter of 19th April, 2022 from the Metro Health Directorate, Tamale (study site) indicating the approval for the conduct of the study at the Metropolis.
- A Completed CHRPE Application Form.
- Participant Information Leaflet and Consent Form.
- Research Protocol.
- Questionnaire.
- Interview Guide and Questionnaire.

The Committee has considered the ethical merit of your submission and approved the protocol. The approval is for a fixed period of one year, beginning **16th August, 2022** to **15th August, 2023** renewable thereafter. The Committee may, however, suspend or withdraw ethical approval at any time if your study is found to contravene the approved protocol.

Data gathered for the study should be used for the approved purposes only. Permission should be sought from the Committee if any amendment to the protocol or use, other than submitted, is made of your research data.

The Committee should be notified of the actual start date of the project and would expect a report on your study, annually or at the close of the project, whichever one comes first. It should also be informed of any publication arising from the study.

Thank you for your application.

Yours faithfully,

Rev. Prof. John Appiah-Poku.
**Honorary Secretary**
**FOR: CHAIRMAN**

Room 7, Block L, School of Medicine and Dentistry, KNUST, University Post Office, Kumasi, Ghana
