## Supplementary material for "Impact of COVID-19 on Antenatal Care Utilization and Health Provider Response in Tamale, Ghana: A Mixed-Method Study": information Sheet and consent form

**PARTICIPANT INFORMATION LEAFLET AND CONSENT FORM**

**Title of Research:**

Effects of COVID-19 on ANC services in the Tamale Metropolitan Area, Tamale.

**Name and affiliation of researcher:**

This is a study conducted by Eunice Addae Azawutor, a student of the Kwame Nkrumah University of Science and Technology (KNUST), school of Public health as part of a partial fulfilment for the award of Master of Science (MSc) in Population, Reproductive and Family Health.

**Background:**

The study’s quest is to find out what the effects of COVID-19 is on antenatal care services in the Tamale Metropolitan Area, Ghana. The consequences of COVID-19 was catastrophic for maternal and newborn health especially for pregnant women (Pant et al., 2020).

**Purpose of the study:**

The purpose of this study is to find out if pregnant women attended the antenatal care clinic the recommended number of times and whether they enjoyed all the services that they are supposed to during the COVID-19 Pandemic in the Tamale Metropolis.

**Procedure of the study, what will be required of each participant and estimated number of participants for the study:**

A total number of 232 participants are estimated to take part in this study. From the four health facilities; Bilpiela health center, Vittin health center, Kpanvo CHPs, and Kotingle CHPs, participants will be selected by systematic random sampling techniques. These selected clients (should they agree to take part in the study), will have to be interviewed. Healthcare service providers who agree to participate will be part of a focus group discussion.

**Risk:**

There are no known anticipated risks in this study apart from the time required for the interview and the focus group discussion. Some of the participants may be embarrassed or uncomfortable in answering some of the questions since they may be sensitive. However, I will humbly request that they respond in a manner that they believe will best express their beliefs on the outlined topics.

**Confidentiality:**

Every information taken for the purpose of this study will be coded. Pieces of information taken in the course of the study cannot be linked to or used to identify any participant. No name or anything that can identify any participant will be used in any publication or reports from this study. Nonetheless, as required by law and as part of the responsibility of this researcher, your records can be made available to the ethics committee as and when required.

**Voluntariness:**

Participating in this study is purely voluntary and therefore at the participant’s free will.

**Alternatives to participation:**

Should anyone choose not to participate in this study, that decision will not affect the quality of care they will receive in this facility or elsewhere.

**Withdrawal from this study:**

You are at liberty to leave the study at any point should you change your mind about participating. You may also choose not to answer any question you find uncomfortable or private.

**Consequence of withdrawal:**

There are no consequences, loss of benefit of care to you at any time of your treatment in this facility or elsewhere should you choose to withdraw from this study. If at the time of withdrawal any non-identifiable information has been modified or used in analysis, reports and publications, they cannot be removed. However, your wishes will be complied with appropriately.

**Cost/Compensation:**

No compensation has been envisaged for participants of this study.

**Contacts:** For further information or inquiry about this study, kindly Contact Eunice Addae Azawutor on **0244715352 or**

Further, if you have any concern about the conduct of this study, your welfare or your rights as a research participant, you may contact:

**The Office of the Chairman**

**Committee on Human Research and Publication Ethics**

**Kumasi**

**CONSENT FORM**

**Statement of person obtaining informed consent:**

I have fully explained this research to ____________________________________ and have given sufficient information about the study, including that on procedures, risks and benefits, to enable the prospective participant make an informed decision to or not to participate.

DATE: _____________________	NAME: _________________________________

**Statement of person giving consent:**

I have read the information on this study/research or have had it translated into a language I understand. I have also talked it over with the interviewer to my satisfaction.

I understand that my participation is voluntary (not compulsory).

I know enough about the purpose, methods, risks and benefits of the research study to decide that I want to take part in it.

I understand that I may freely stop being part of this study at any time without having to explain myself.

I have received a copy of this information leaflet and consent form to keep for myself.

NAME:_________________________________________________________________

DATE: ____________	SIGNATURE/THUMB PRINT: ___________________

**Statement of person witnessing consent (Process for Non-Literate Participants):**

I	(Name of Witness) certify that information given to

(Name of Participant), in the local language, is a true reflection of what l have read from the study Participant Information Leaflet, attached.

WITNESS’ SIGNATURE (maintain if participant is non-literate): ____________________

MOTHER’S SIGNATURE (maintain if participant is under 18 years): ________________

MOTHER’S NAME: ______________________________________________________

FATHER’S SIGNATURE (maintain if participant is under 18 years): _________________

FATHER’S NAME: ______________________________________________________
